# Case fatality rate of novel coronavirus disease 2019 in China

**DOI:** 10.1101/2020.02.26.20028076

**Authors:** Rui Qi, Chao Ye, Xiang-rong Qin, Xue-Jie Yu

## Abstract

**Background:** A pandemic of coronavirus disease 2019 (COVID-19) which have caused more than 80 thousand persons infected globally is still ongoing. This study aims to calculate its case fatality rate (CFR).

**Methods:** The method, termed as converged CFR calculation, was based on the formula of dividing the number of known deaths by the number of confirmed cases T days before, where T was an average time period from case confirmation to death. It was found that supposing a T, if it was smaller (bigger) than the true T, calculated CFRs would gradually increase (decrease) to infinitely near the true T with time went on. According to the law, the true T value could be determined by trends of daily CFRs calculated with different assumed T values (left of true T is decreasing, right is increasing). Then the CFR could be calculated.

**Results:** CFR of COVID-19 in China except Hubei Province was 0.8% to 0.9%. So far, the CFR had accurately predicted the death numbers more than 3 weeks. CFR in Hubei of China was 5.4% by which the calculated death number corresponded with the reported number for 2 weeks.

**Conclusion:** The method could be used for CFR calculating while pandemics are still ongoing. Dynamic monitoring of the daily CFRs trends could help outbreak-controller to have a clear vision in the timeliness of the case confirmation.

## Introduction

An outbreak of pneumonia caused by a novel coronavirus occurred in Wuhan, Hubei Province, China at the end of 2019.(1) On Feb 11, 2020 the World Health Organization (WHO) announced an official name for the disease as coronavirus disease 2019 (COVID-19) and the International Committee on Taxonomy of Viruses named the novel coronavirus as severe acute respiratory syndrome coronavirus 2 (SARS-CoV-2). The outbreak of the disease was linked to a live animal market firstly and then was reported person-to-person transmission.(2, 3) The disease has rapidly spread from Wuhan City to other areas. As of Mar 1, 2020, approximately 80 thousand cases in China alone have been confirmed. Cases also have been reported in more than 25 countries of 5 continents. The case fatality rate (CFR) represents the proportion of people who eventually die from a specified disease. CFR typically is used as a measure of disease severity and is often used for prognosis where comparatively high rates are indicative of relatively poor outcomes (4). It also can be used to compare the effect of treatments among different areas. In general, when a pandemic has ended, CFR can be calculated by dividing the number of known deaths by the number of confirmed cases. A major difficulty in estimating case fatality rate is ensuring the accuracy of the numerator and the denominator. While a pandemic is still ongoing, it is tempting to estimate the case fatality rate by dividing the number of known deaths by the number of confirmed cases reported so far. The resulting number, called naive CFR, however, does not represent the true case fatality rate because this calculation does not account for the delay between case confirmation and disease outcome (5). In that case, the CFR will be underestimated. To estimate the CFR while a pandemic is still ongoing, the denominator should be corrected as cases at T days before, where T is an average time period from case confirmation to death. This study aims to calculate the CFR of the COVID-19 in China by estimating the average time period from case confirmation to death.

## Methods

Data: population level data in this study included daily accumulative numbers of cases and deaths of COVID-19 in China from Jan 21 to Mar 1, 2020. Data was collected from National Health Commission of China, China CDC, and provincial level health authorities.

Estimation of T (average time period from case confirmation to death): To calculate CFR, it should be realized that deaths at day X are averagely from cases at day X-T rather than day X. Given a T value, a group of CFRs (daily CFRs) can be obtained from different X days. As known that death number at day X should be less than case number at day X-T (if more than day X-T, CFR would be greater than 100% which is illogical). Based on this point, the range of T can be narrowed. More importantly, no matter what T value is assumed, even it is far away from the true T value, the daily CFRs would converge towards (infinitely approach to but never be over) the true CFR with time (X) increases. The following example will illustrate this principle (Table 1). Assuming CFR = 10%, T = 4 for a disease, the cases number was from 100 to 10000 at day X (X=1 to 100), then the deaths number would be 10 (10, 20 and so on) at day X+4 (5, 6 and so on). When calculating daily CFRs based on case and death numbers with formula deaths (X) divided by cases (X-T), law 1: if assumed T was equal to the true T value (4 in the example), calculated daily CFRs at different day X would constantly be the true CFR (0.1); if assumed T was greater than the true T (5 and 6), daily CFRs would be greater than the true CFR (0.1) and infinitely reduce to near it with the time (X) increased; if assumed T was smaller than the true T (1 to 3), daily CFRs would be smaller than the true CFR and infinitely increase to near it. Besides, it could be found that, law 2: if assumed T was more far away (bigger absolute difference) from the true T, daily CFRs would be more far away from the true CFR and they would need more times to converge towards it. In this example, case numbers were given from 100 to 10000 by 100 increments per day, however, cases growth every day would not be evenly for an infectious disease. Then case numbers in this example were replaced by real case numbers of COVID-19 and the convergence tendency still remained except for individual data points. Based on convergence laws, we used exhaustive method to calculate daily CFRs of COVID-19 by different T values. If an assumed T resulted in relatively constant daily CFRs, and T+1 resulted in decreasing daily CFRs and T-1 in increasing, it could be determined as the true T. The method above could be termed as converged CFR calculation.

**Table 1.**
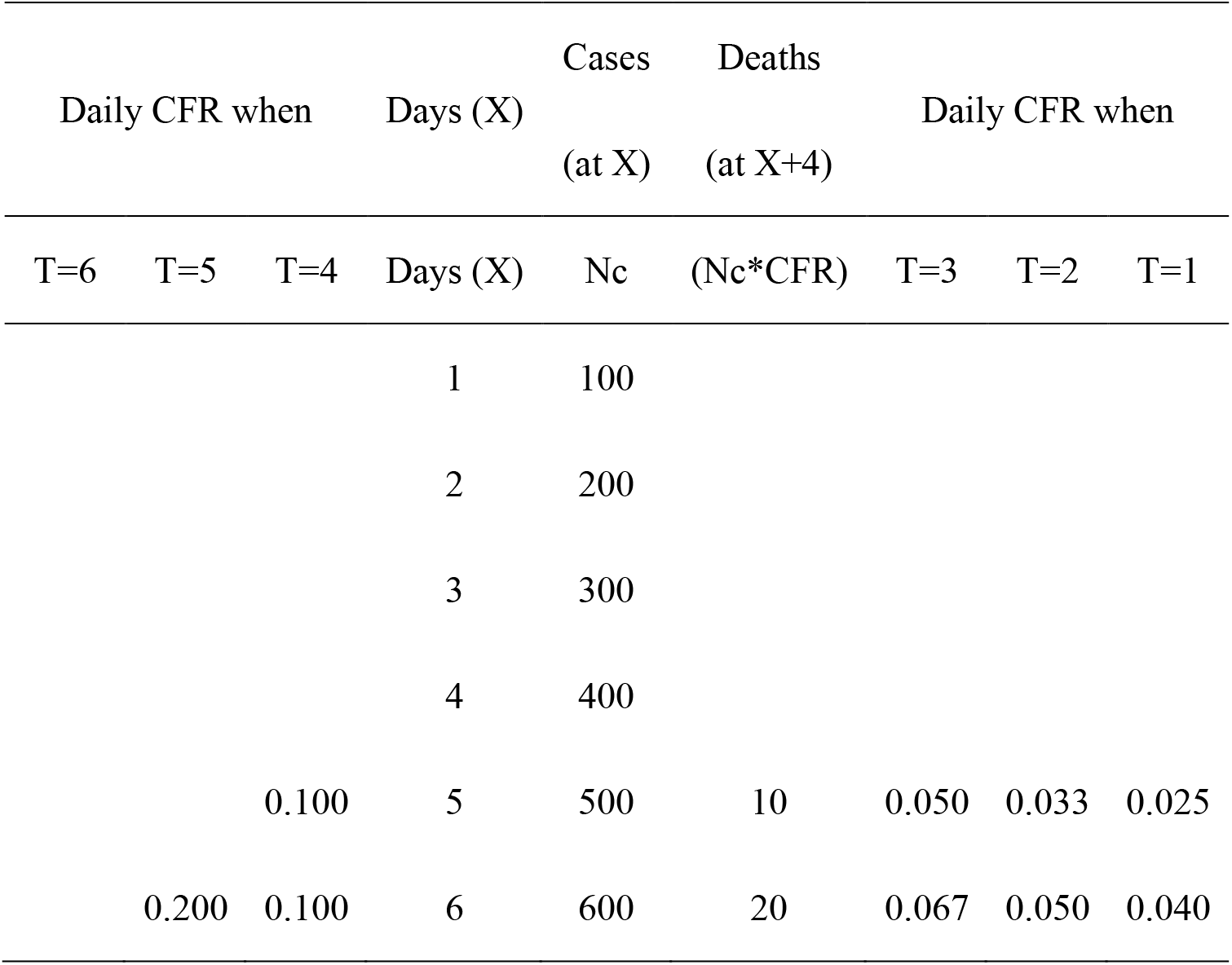
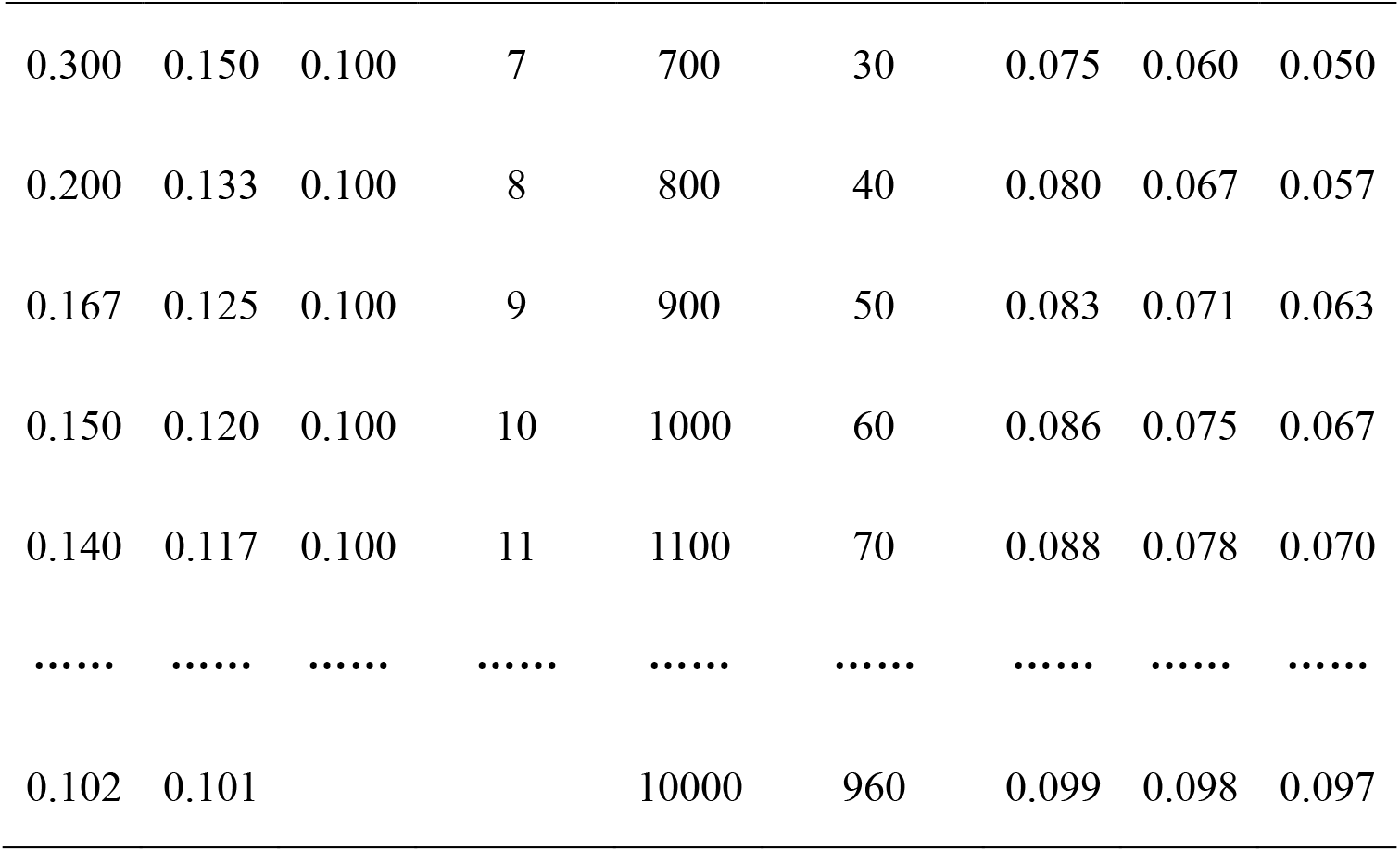
Convergence tendency at different T values for a CFR=10% assumed disease

## Results

### CFR in China except Hubei Province (non-Hubei regions)

A number of T values were selected for screening based on convergence laws. After different T values were tried, as Figure 1 showed, when assumed T was 11, the daily CFRs were decreasing and had no pronounced increase, when it was 0 to 7; the daily CFRs had pronounced increase after early time (T > 11 were not shown due to continuously decreasing trends). CFRs increased as expected according to laws at later stage in some assumed T values (e.g. T=0), but it decreased at early stage which seemed not satisfy the convergence laws. Actually, it was normal. Convergence laws happened due to the force of the true CFR drawing daily CFRs towards its direction by dominating accumulated death numbers. At early stage, the outcome of death had not yet occurred resulting in daily CFRs decreasing with the growth of case number. Thus, T value exploration by convergence laws should depend on period of death growth.

**Figure 1.**
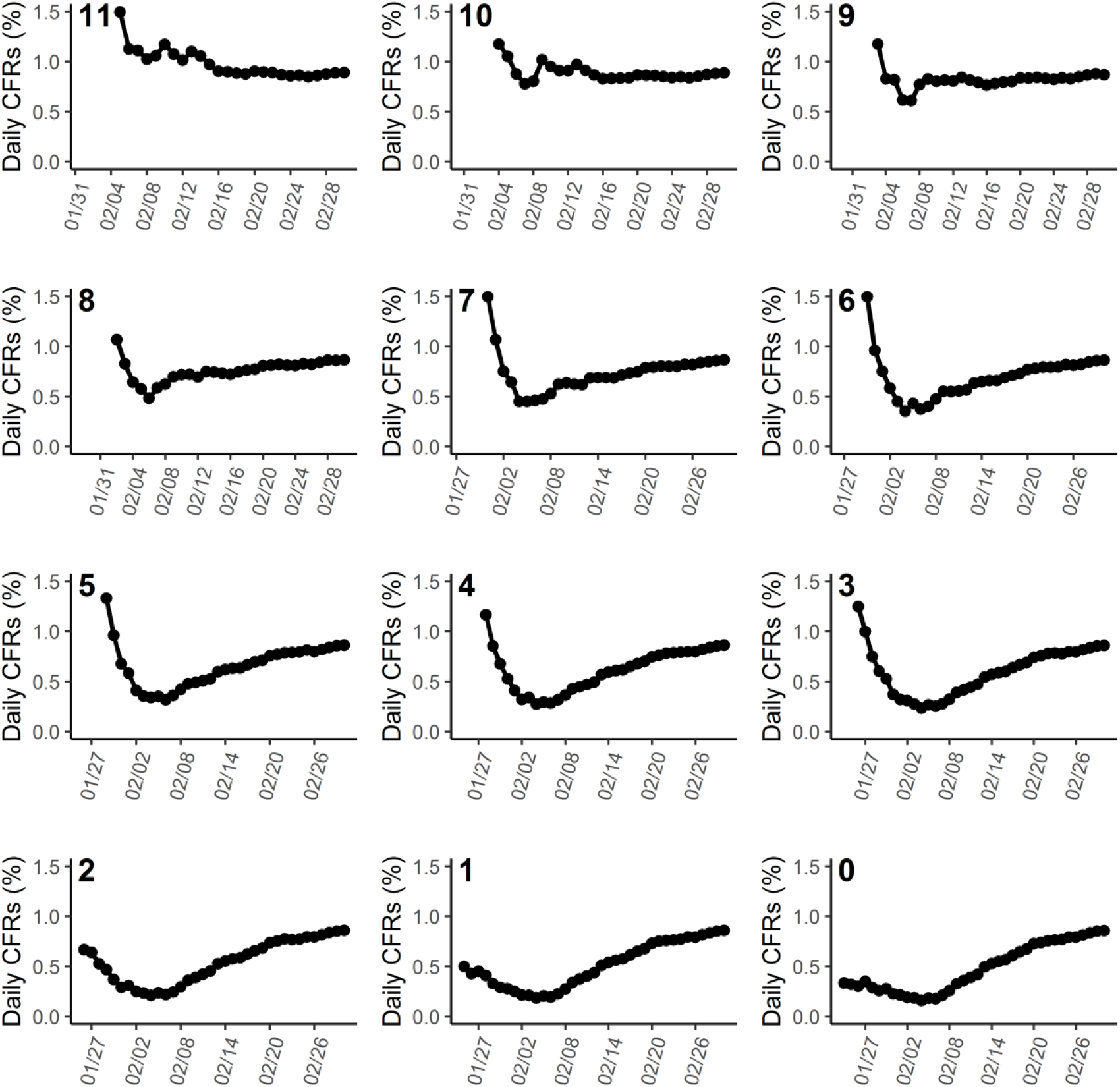
Calculated daily CFRs of non-Hubei regions by Mar 1 when T was assumed from 0 to 11.

Results of Figure 1 indicated the true T should be in the range of 8 to 10. As differences between CFRs were too small at converging stage to compare and scales of y axis in different plots of Figure 1 varied greatly, Figure 1 was only used for preliminary tendency exploration. Converging stage CFRs had been cut out to plot with same y axis scales for the true T and CFR estimation (Figure 2).

**Figure 2.**
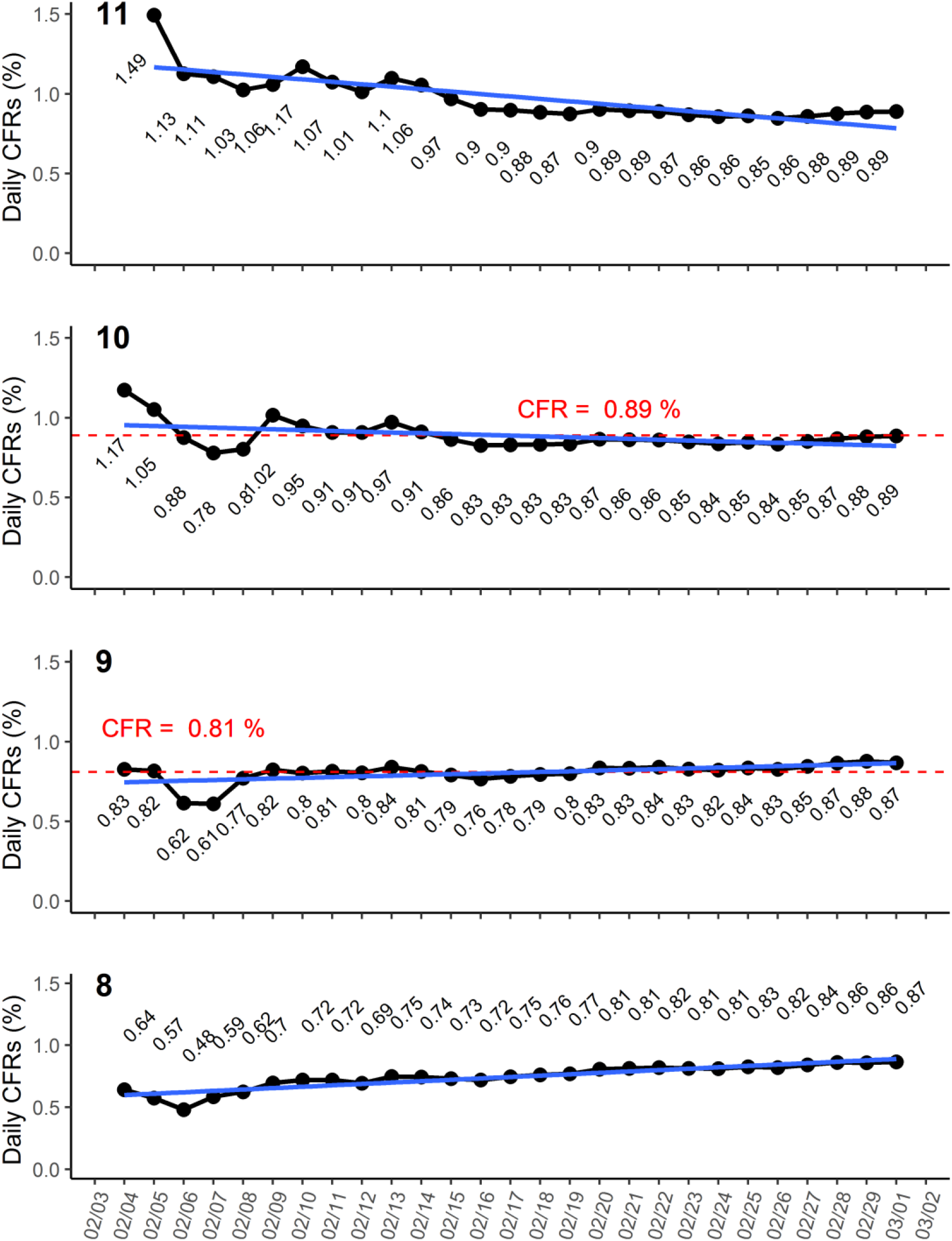
Converging stage daily CFRs of non-Hubei regions when T was from 8 to 11 Blue lines represented linear models generated for analysis of variances and linear trends of these data points in plots. Red dotted line and number were the estimated true CFR.

As mentioned in Methods, with the time increased, even under a false T in calculation, the daily CFR could converge towards the true CFR though more times needed. If assumed T was equal to the true T value, calculated daily CFRs would keep constant. As Figure 2 showed, for T= 11 and 8, comparing with T = 9 and 10, CFRs still had slightly decreasing and increasing trends, respectively. Linear models (blue lines) were generated for analysis of variances and linear trends of theses CFR points in each plot. The slopes of models became flatter and approached towards to 0 when T was from 8 to 9 and 11 to 10. The results indicated the true T should be bigger than 8 and less than 11. When T = 9, the CFRs were almost staying in one line (red dotted line in Figure 2) and slightly increased later. When T = 10, though the daily CFRs decreased early but quickly they reached a stable stage. So the true T might be between 9 and 10 days. The mean values of data in plot 9 and 10 of Figure 2 were 0.8% and 0.9%, respectively. The true CFR of COVID-19 in China except Hubei Province should fall between 0.8% and 0.9%. An assumed T was the closer to the true T value, the earlier daily CFRs converging to the true CFR happened. The mean value of CFRs at later stage of plot 9 or 10 was approximately 0.85%. As shown in Figure 2, if data was analyzed before Feb 20, T = 9 (0.81%) might be determined as the true T value (true CFR). But now the T was postponed to between 9 and 10 days. The reason was not the uncertainty of method, but the long disease course of COVID 19. Time of case confirmation to outcome was longer in some cases than most which caused the true T bigger and CFR slightly increased. For non-Hubei regions, on Feb 20, 6719 of 12644 (53%) cases were in hospitals, but on Mar 1, only 2461 of 13061 (19%) were in hospitals. When a pandemic ended, the naive CFR could be equal to the true CFR. The COVID 19 epidemic in non-Hubei regions is coming to the end (only sporadic cases were reported recently in the non-Hubei regions), so far, the naive CFR was 0.85% (112/13061). The final CFR should not be bigger too much than 0.85%. It indicated calculated CFR by this converged CFR calculation method was a good estimation of the true CFR. More importantly, it could be approximately estimated earlier (3 weeks ago) by our method when a pandemic was still ongoing.

### CFR in Hubei Province

As shown in Table 2, after Feb 3, death number (350) were more than Jan 21 case number (270), if the T was 12 (Feb 3 minus Jan 1), the CFR would be illogically greater than 100%. In another words, death numbers only when before day 12 were less than case number at day 1. So the time T should be less than or equal to 11 days (12-1). The death number when was firstly more than the case number at day 2 was Feb 5 (day 15), so the T should be less than or equal to 12 (14-2). The rest could be done in the same manner. Finally, the smallest T value (T = 11) was selected as the upper limit for convergence screening.

**Table 2.**
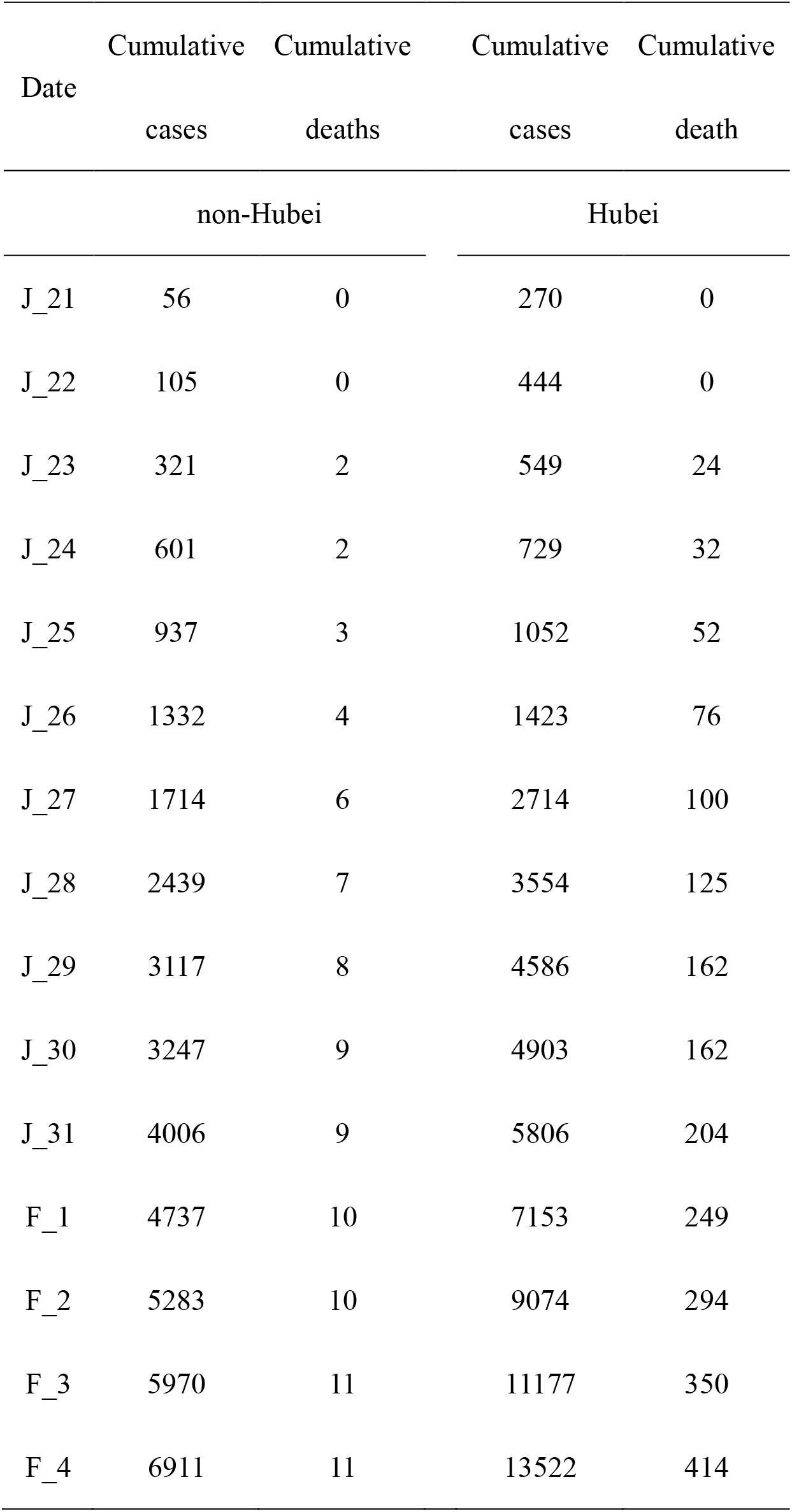
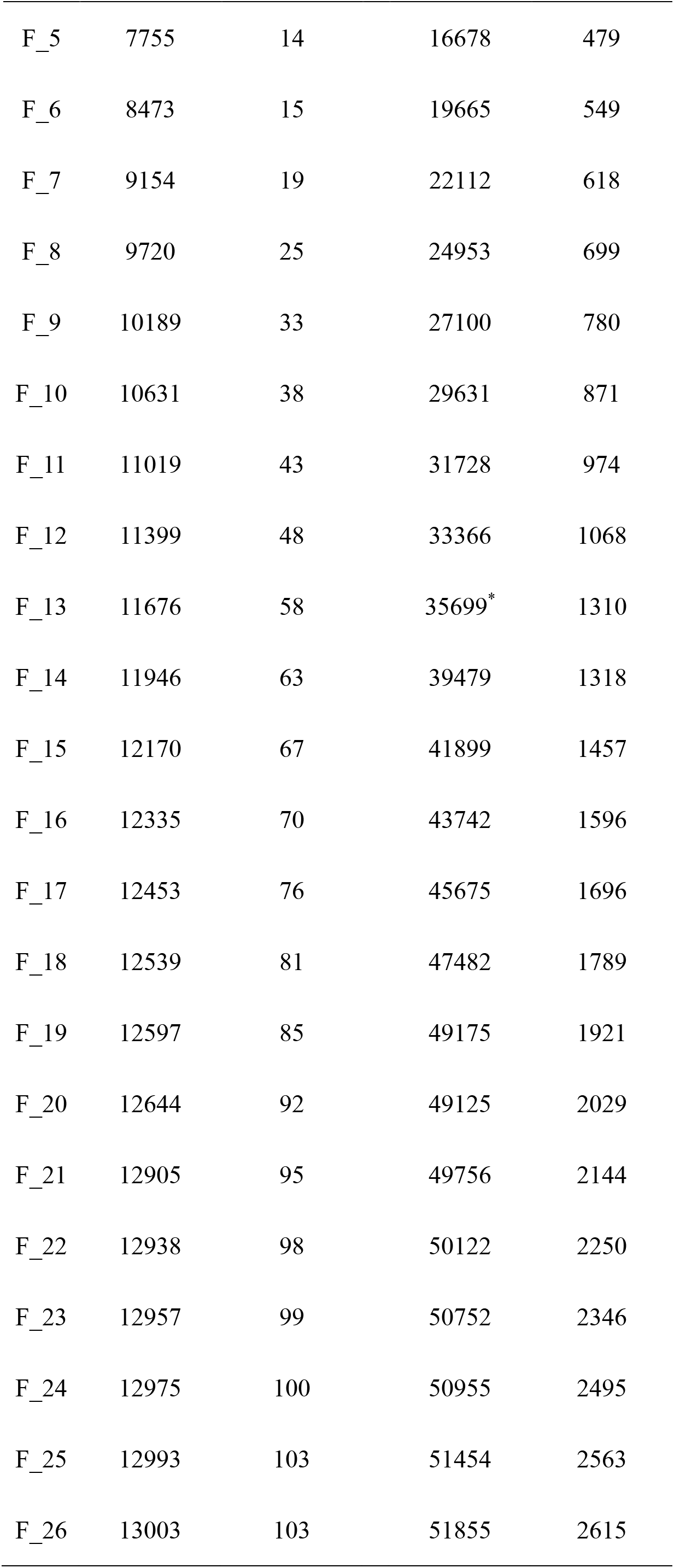
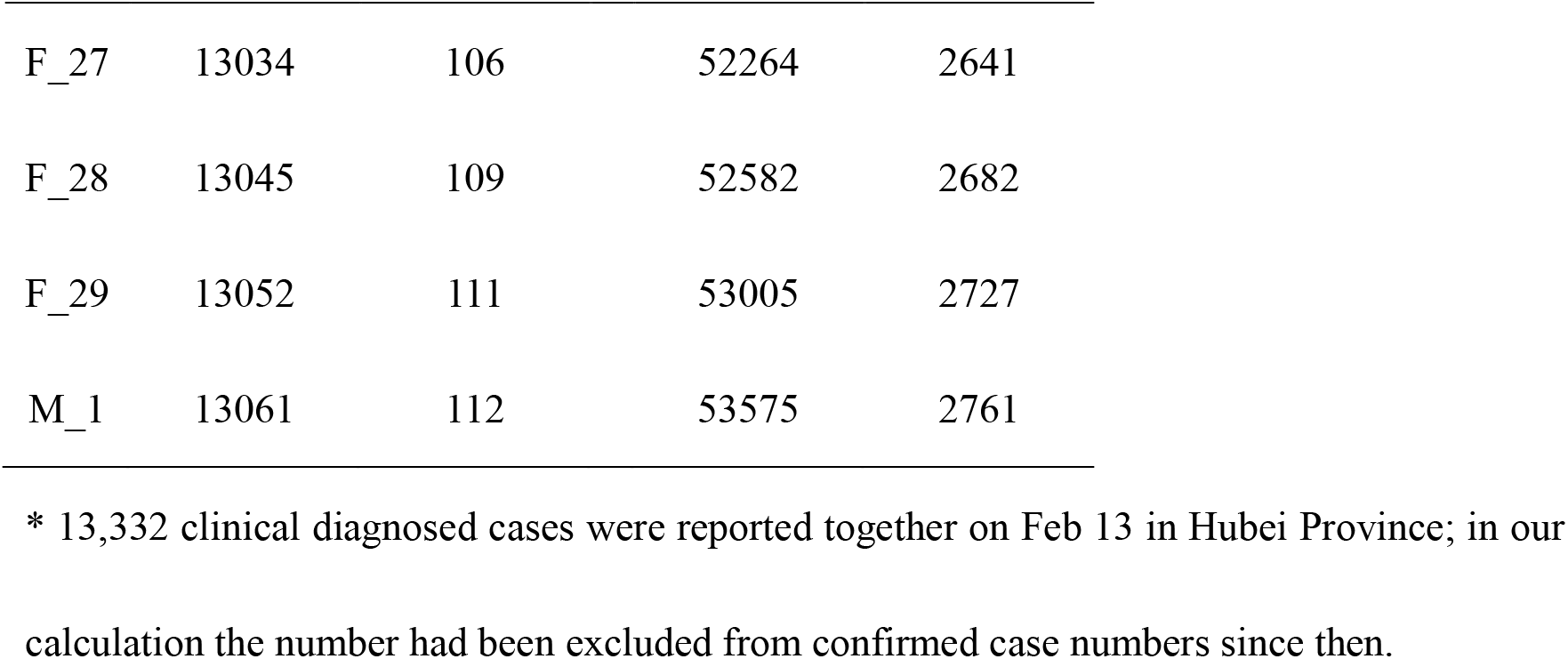
Death and case numbers of COVID-19 in China from Jan 21 to Mar 1

| Date | Cumulative<br>cases | Cumulative<br>deaths | Cumulative<br>cases | Cumulative<br>death |
| --- | --- | --- | --- | --- |
|  | non-Hubei |  | Hubei |  |
| J_21 | 56 | 0 | 270 | 0 |
| J_22 | 105 | 0 | 444 | 0 |
| J_23 | 321 | 2 | 549 | 24 |
| J_24 | 601 | 2 | 729 | 32 |
| J_25 | 937 | 3 | 1052 | 52 |
| J_26 | 1332 | 4 | 1423 | 76 |
| J_27 | 1714 | 6 | 2714 | 100 |
| J_28 | 2439 | 7 | 3554 | 125 |
| J_29 | 3117 | 8 | 4586 | 162 |
| J_30 | 3247 | 9 | 4903 | 162 |
| J_31 | 4006 | 9 | 5806 | 204 |
| F_1 | 4737 | 10 | 7153 | 249 |
| F_2 | 5283 | 10 | 9074 | 294 |
| F_3 | 5970 | 11 | 11177 | 350 |
| F_4 | 6911 | 11 | 13522 | 414 |
| F_5 | 7755 | 14 | 16678 | 479 |
| F_6 | 8473 | 15 | 19665 | 549 |
| F_7 | 9154 | 19 | 22112 | 618 |
| F_8 | 9720 | 25 | 24953 | 699 |
| F_9 | 10189 | 33 | 27100 | 780 |
| F_10 | 10631 | 38 | 29631 | 871 |
| F_11 | 11019 | 43 | 31728 | 974 |
| F_12 | 11399 | 48 | 33366 | 1068 |
| F_13 | 11676 | 58 | 35699* | 1310 |
| F_14 | 11946 | 63 | 39479 | 1318 |
| F_15 | 12170 | 67 | 41899 | 1457 |
| F_16 | 12335 | 70 | 43742 | 1596 |
| F_17 | 12453 | 76 | 45675 | 1696 |
| F_18 | 12539 | 81 | 47482 | 1789 |
| F_19 | 12597 | 85 | 49175 | 1921 |
| F_20 | 12644 | 92 | 49125 | 2029 |
| F_21 | 12905 | 95 | 49756 | 2144 |
| F_22 | 12938 | 98 | 50122 | 2250 |
| F_23 | 12957 | 99 | 50752 | 2346 |
| F_24 | 12975 | 100 | 50955 | 2495 |
| F_25 | 12993 | 103 | 51454 | 2563 |
| F_26 | 13003 | 103 | 51855 | 2615 |
| F_27 | 13034 | 106 | 52264 | 2641 |
| F_28 | 13045 | 109 | 52582 | 2682 |
| F_29 | 13052 | 111 | 53005 | 2727 |
| M_1 | 13061 | 112 | 53575 | 2761 |

\* 13,332 clinical diagnosed cases were reported together on Feb 13 in Hubei Province; in our calculation the number had been excluded from confirmed case numbers since then.

Figure 3 was the calculation of daily CFRs with assumed T values (0 to 11). When assumed T was 8 to 11, daily CFRs were continuously decreasing. When T = 0 and 3, there were increase trends at later stage which meant they were smaller than the true T value. Converging stage CFRs data when T = 4 to 7 was selected for plotting with the same y axis scales (Figure 4). As it showed, For T= 4, CFRs had increase trends, and T = 5, the CFRs slightly increased. When T was 7, CFRs decreased and reached stable at later stage. When T was 6, plateau stage appeared earlier than T = 7. The slopes of linear models became flatter and approached towards to 0 when T approaching to 6. Then T = 6 was selected as the true T value for the true CFR calculation. The true CFR of COVID-19 in Hubei calculated by mean value of the daily CFRs of plot 6 in Figure 4 was 5.4%. The estimated T value was smaller than non-Hubei regions. It was not surprising as it seemed that time of case confirmation to death was shorter. Previously in Wuhan City of Hubei Province, many patients had not been confirmed and reported timely due to overwhelmed medical services and lack of testing kits. The death number (from confirmed and unconfirmed population) could prefer to “select” forward case pools with bigger population. Thus, to obtain an accurate CFR, timeliness of case conformation should not vary too much. The possibility could not be rule out that the CFR might slightly increase later like non-Hubei regions due to the long disease course of COVID 19.

**Figure 3.**
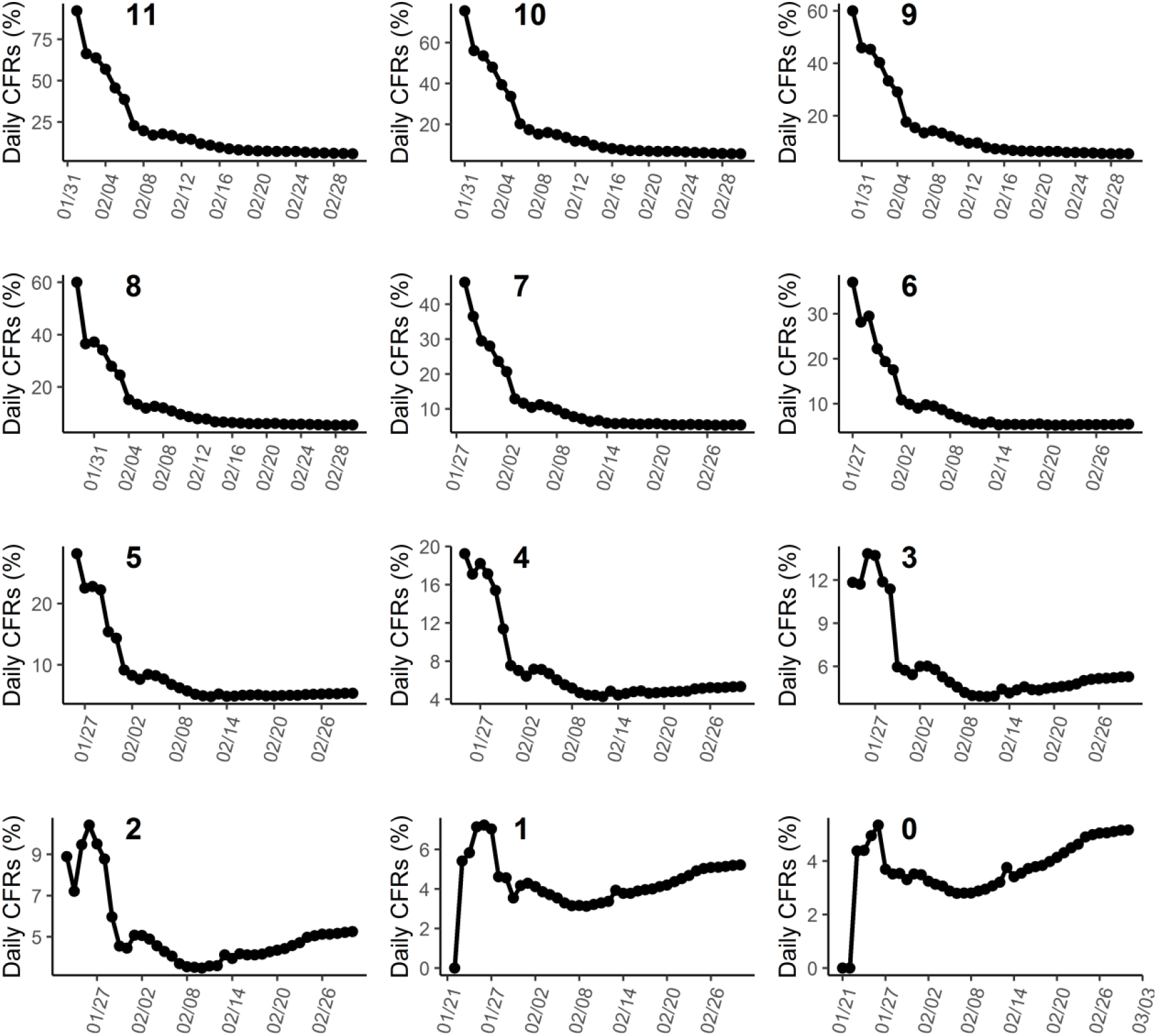
Calculated daily CFRs of Hubei Province by Mar 1 when T was from 0 to 11.

**Figure 4.**
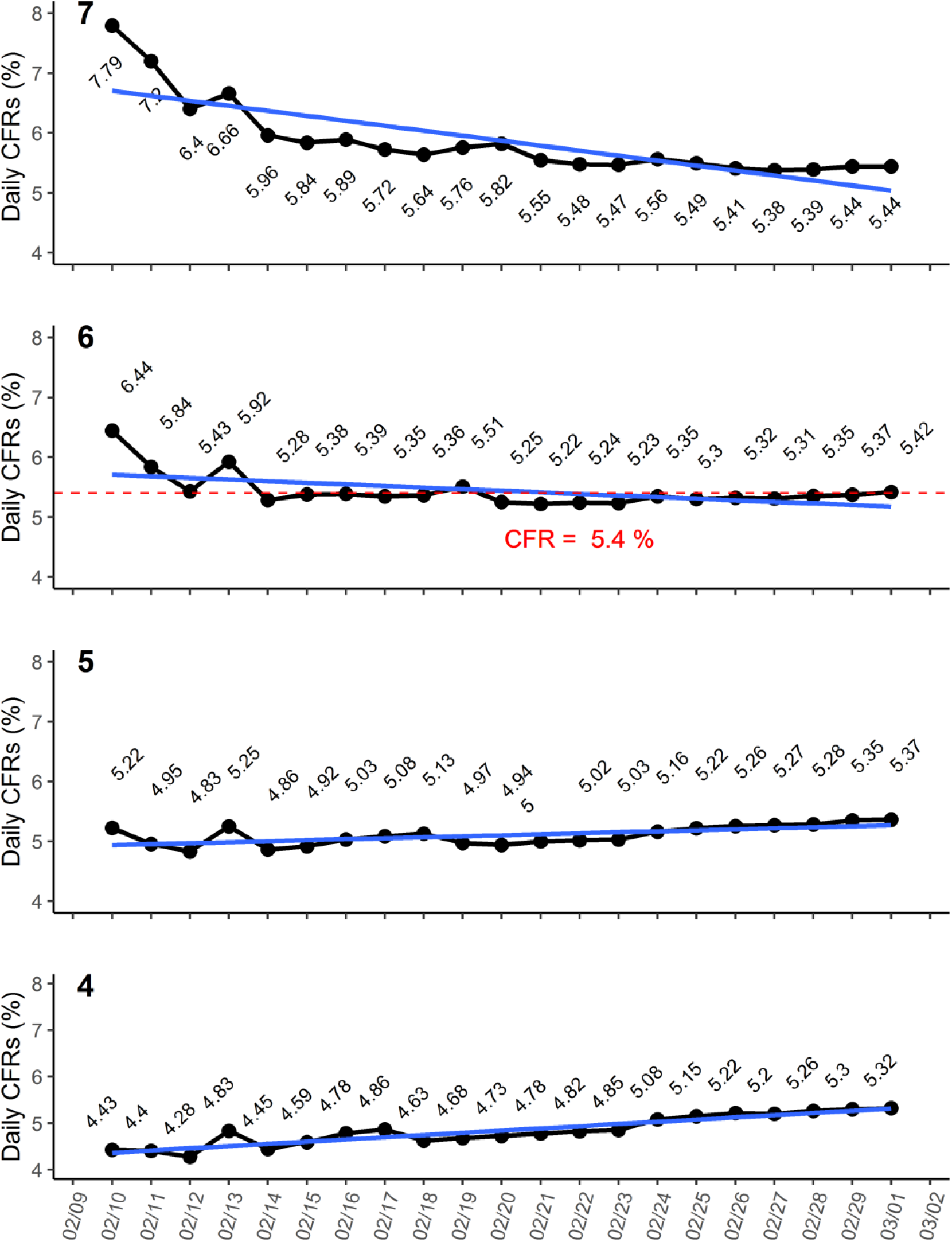
Converging stage daily CFRs of Hubei Province when T was from 4 to 7 Blue lines represented linear models generated for analysis of variances and linear trends of these data points in plots. Red dotted line and number were the true CFR.

### Validation of calculation

True numbers of death were compared with numbers estimated by the calculated T and CFR to validate the accuracy of our method. The cumulative cases at day X multiplied calculated CFR should be approximately equal to true death number at day X+T theoretically. As shown in Figure 5-non-Hubei, since Feb 4, calculated death numbers had a good fit to the true death data. The curves came closest to coinciding in shape. For Hubei (Figure 5-Hubei), the predictive curve was similar in shape with true death line, however, from Jan 23 to Feb 10, predicted death numbers were smaller than the true numbers. The predicted curve from Jan 23 to Feb 10 seemed be moved to right about 2 or 3 days. A subset data from Jan 21 to Feb 12 was selected to recalculate the T, and results in Hubei, T was 2 days. However, it could be found in plot 2 of Figure 3 that only 3 points stayed in the stable line before Feb 12. Without later data, it could result in a misleading false CFR. Thus, for CFR calculation, the stable stage of daily CFRs should not be too short.

**Figure 5.**
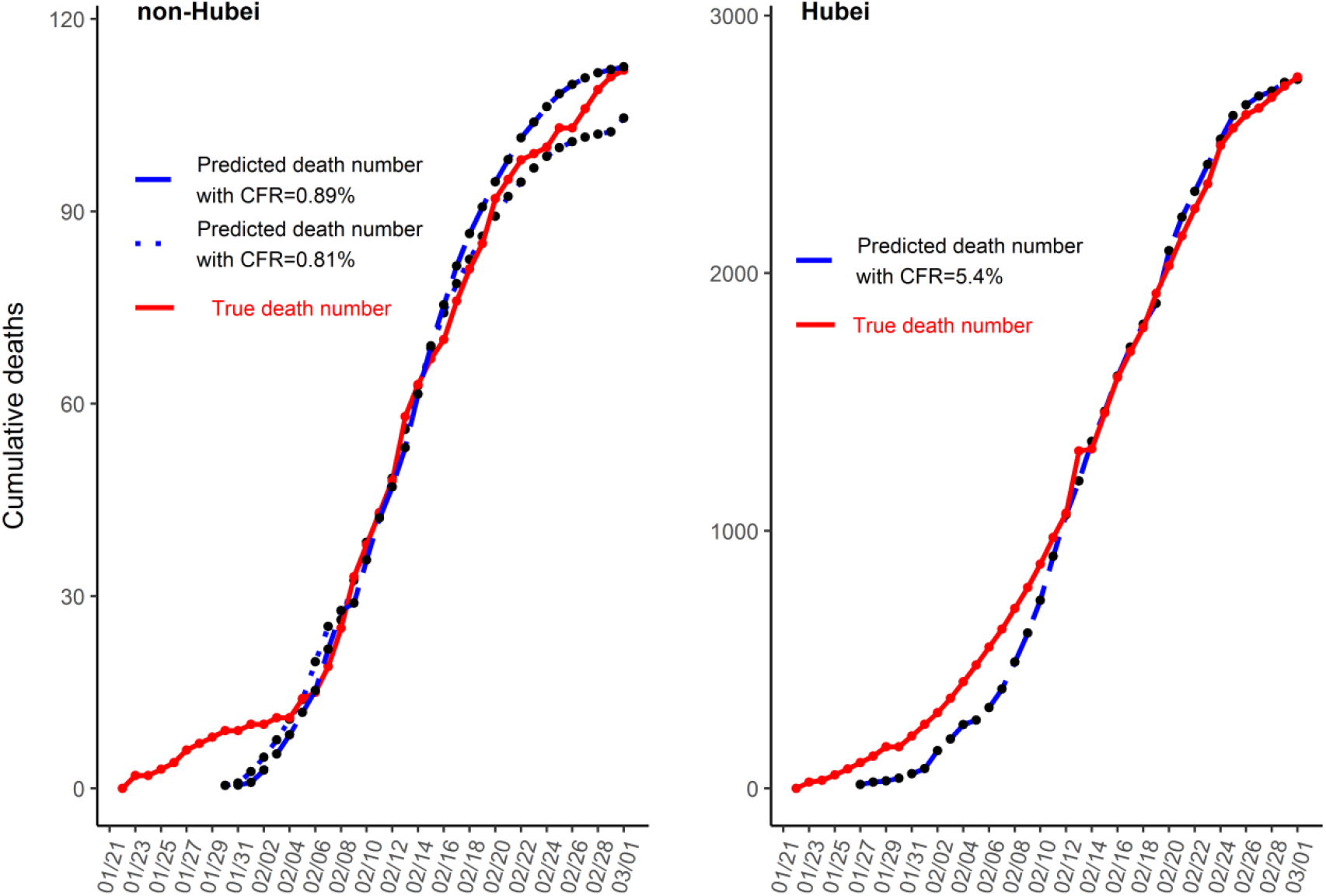
Comparison of true numbers of death and estimated numbers by the calculated T and CFR.

On the other hand, T became bigger indicated the case confirmation was timelier. Outbreak-controllers could indirectly have information about timeliness of case confirmation by monitoring daily CFRs. Stable CFRs trends meant the denominator for CFR calculation, case number, was accurate enough. As extant cases were in quarantine, combining with transmission potential of diseases, it could provide policy-makers information about the risk of second infection, which could help them with evaluation of when people in regions could go back on production. In summary, as death numbers had been almost accurately predicated by calculated true CFR for more than 3 weeks, it could be considered as the true CFR of COVID-19 in China except Hubei Province. For Hubei, calculated death number corresponded with the reported number for more than 2 weeks.

## Discussion

CFR was calculated by dividing the number of deaths from a specified disease. For a infectious disease, the outcome of death were determined by virulence of causative pathogens, immunity and health status of those infected, medical conditions, received treatment and so on. Whether all infected cases had been completely included into the denominator also affected the CFR. That meant, for the same disease, CFR were not always constant and could vary between populations (6). COVID-19 firstly occurred in Wuhan City, Hubei Province, China and quickly went into a big outbreak and overwhelmed local medical facilities. Then it extended to the whole Hubei Province and other regions in China during the heavy-travel Chinese Spring Festival holidays. The Chinese government rapidly isolated Wuhan and took emergency measure nationwide to prevent and control disease. Non-Hubei regions response to COVID-19 could be regarded as timely. The situations of outbreak in Hubei and non-Hubei regions were quite different. So CFRs were calculated separately. Diagnose and confirmation towards patients presenting with more severe disease had priority in Hubei, especially Wuhan as the limited healthcare-facilities and testing capacities. Thus, the calculated CFR for Hubei was higher due to the underdetection of mild or asymptomatic cases. Other regions in China had token completely epidemical investigation of diagnosed cases under the nationwide strict quarantine and screening policy. Close contactor investigation by CDC could help find mild or asymptomatic cases. Thus, CFR calculated from these regions could be regarded as accurate values in the situation of medical services were not overwhelmed.

So far, only a few studies reported CFR of COVID-19. Study of Wuhan’s earliest 41 cases gave a 15% death rate (7). However, regardless of the sample size, these cases were highly biased towards the more severe cases for CFR calculation. Another study reported the CFR was 4.3% which also had a biased study population (Wuhan hospitalized patients) (8). A newly epidemiological study estimated the CFR was 3.06% (95% CI 2.02-4.59%) from 4,021 cases (9). This study included data from non-Hubei regions, so the CFR should be smaller than that of Wuhan. When epidemic was still ongoing, CFR could be estimated by following a cohort, however, it was time-consuming and difficult to included size-enough and representative patients from unbiased population. Considering the features of daily CFRs convergence, true CFR estimation based on population-level big data might be a good way.

In our study, calculated T values were different, T was between 9 and 10 in non-Hubei but was 6 in Hubei. The time in Hubei from confirmation to death was shorter comparing with non-Hubei. On Feb 13, more than 10 thousands cases were reported one day including clinical diagnosed cases without laboratory confirmation. It indicated there would be a lag in case confirmation in Hubei. With the cases in Hubei were confirmed timely, the estimated T might move towards bigger. In addition, when the factors causing the true CFR higher in Hubei were controlled, the true CFR might decrease. The daily CFRs in Figure 3 would also change itself to the new T and CFR. But, 5.4% was the true CFR at the present stage. In our study, 13332 clinical cases were excluded since Feb 13, which might result in an overestimation of the true CFR.

CFR calculated in our study was dynamic, which could be used to real-time monitor the case confirmation situation. If daily CFRs kept on a horizontal line and the confirmed cases were continuously decreasing, it meant the control measures had worked well. Not only infectious diseases, but also other diseases which were difficult to follow cohorts can be monitored to calculate the CFR. But a limitation should be taken into consideration that daily CFR would approach to true CFR only when deaths started to appear. When calculated T was too small, it might give outbreak-controllers information that if there remained a lot of infected persons unconfirmed. And if calculated T started to move backwards with time, it meant confirmation of patients had become timely at then.

In conclusion, by converged CFR calculation method, the true CFR of COVID-19 in China except Hubei Province was approximately from 0.8% to 0.9%. This calculated CFR could accurately predict the death numbers for more than 3 weeks. The CFR in Huibei was 5.4% at the present stage. This method in our study can be used for CFR calculation when a pandemic is still ongoing and monitoring the case confirmation situation.

## Data Availability

All data was avaliable in the manuscript

## Acknowledgments

This study was partially supported by a grant from National Natural Science Funds of China (No. 31570167).

